# Diagnostic Strategies to Identify Patients with Genetic Salt-Losing Tubulopathies

**DOI:** 10.1101/2021.12.12.21267676

**Authors:** Elizabeth Wan, Daniela Iancu, Emma Ashton, Keith Siew, Barian Mohidin, Chih-Chien Sung, China Nagano, Detlef Bockenhauer, Shih-Hua Lin, Kandai Nozu, Stephen B Walsh

## Abstract

**Background:** Distinguishing patients with the inherited salt-losing tubulopathies (SLT), Gitelman or Bartter syndrome (GS or BS) from wildtype (WT) patients who purge is difficult. We decided to identify clinical/biochemical characteristics which correctly classify SLT.

**Methods:** 66 patients with possible SLT were recruited to a prospective observational cohort study at the UCL Renal Tubular Clinic (London). 31 datapoints were recorded on each patient. All patients were genotyped for pathogenic mutations in genes which cause SLT; 39 patients had pathogenic variants in genes causing SLT. We obtained similar datasets from cohorts in Taipei and Kobe; the combined dataset comprised 419 patients, 291 had genetically confirmed SLT. London and Taipei datasets were combined to train machine learning (ML) algorithms. These were then tested on the Kobe dataset to determine the best biochemical predictors of genetic confirmation of SLT.

**Results:** Single biochemical variables (e.g. plasma renin) were significantly, but inconsistently different between SLT and WT, in the London and combined cohorts.

A decision table algorithm using serum bicarbonate and urinary sodium excretion (FE_Na_) achieved a classification accuracy of 74%. A simpler algorithm based on the FE_Cl_ achieved a classification accuracy of 61%. This was superior to all of the single biochemical variables identified previously.

## Introduction

Chronic hypokalaemia due to renal wasting, associated with a metabolic alkalosis but without hypertension is a frequent presentation of Gitelman syndrome (GS) and classic, type 3, Bartter syndrome (BS3), which are hereditary salt-losing tubulopathies (SLT).

GS is caused by inactivating mutations of *SLC12A3*, which encodes the apical thiazide-sensitive sodium chloride cotransporter (NCC) of the distal convoluted tubule (DCT)^1^. It characteristically causes a hypokalaemic metabolic alkalosis with hypomagnesaemia and hypocalciuria, although hypomagnesaemia may be absent and the hypocalciuria is very variable^2^. It is an autosomal recessive disease with an estimated mutant gene frequency of between 1-3.6%^3^. Its overall prevalence is ∼10/40 000, making it the most common inherited tubulopathy^4^.

Bartter syndrome (BS) is also characterised by hypokalaemic metabolic alkalosis. This is due to underactivity of the bumetanide-sensitive sodium potassium chloride cotransporter, (NKCC2) present in the thick ascending limb (TAL) of the loop of Henle. Types 1, 2 and 4 Bartter syndromes are associated with severe salt and water loss and failure to thrive in infancy and are rarely diagnosed in adults. BS3 is the commonest form and is caused by mutations in *CLCNKB*, which encodes a basolateral chloride channel, CLCKb^5^ . Like GS, it is often diagnosed following the incidental discovery of hypokalaemia and alkalosis in adulthood. Although the Bartter syndrome complex tends to cause hypercalciuria and not hypomagnesaemia, BS3 has a variable phenotype that may be indistinguishable from Gitelman. This is probably due to the distribution of CLCKb which is present both in the TAL and

DCT^6^. This has led some authors to suggest reclassifying GS and BS on the basis of the location of the lesion, either in the TAL or DCT^6^.

The clinical diagnosis of GS and BS is made more difficult by conditions which mimic their biochemical phenotype (a hypokalaemic metabolic alkalosis). In children, it may be mimicked by inherited conditions, e.g. congenital chloride diarrhoea or cystic fibrosis. In adults, it can be mimicked by vomiting, laxative abuse or surreptitious diuretic use^7^. Together these purging behaviours have been called ‘Pseudo-Bartter’ syndrome, and in some series comprise over 50% of patients being investigated for hypokalaemia^8^. As they are almost always surreptitious and indicative of an often-undiagnosed eating disorder, they can be difficult to distinguish from either GS or BS without diagnostic genetic testing.

However, it is important to diagnose the ‘pseudo-Bartter’ purging behaviours. Purging patients are likely to take up time and resources in a clinical environment (a nephrology clinic) that is inappropriate for them. Further, diagnosing a purging behaviour is significant as it is likely to be a presentation of bulimia nervosa, or another eating disorder. Patients with bulimia have an all-cause mortality of up to 8 times that of the general population^9^ and 3 times greater than that for bipolar disorder and depression^10^. An early diagnosis and referral to appropriate psychiatric services may be lifesaving.

It is therefore unsurprising that genetic testing is recommended for diagnosis by the KDIGO working group on Gitelman syndrome^11^. However, even now, access to genetic testing is unequal in centres across the UK, and the disparity is greater in many other countries. We describe a well characterised cohort of patients who were investigated for possible SLT by genetic and biochemical testing. We combine this data with 2 other similar datasets and apply statistical and machine learning approaches to identify strategies to predict which patients will have biallelic mutations causing GS or BS.

## Materials and Methods

The UCL Department of Renal Medicine runs a specialist clinic for renal tubular disorders. A significant number of patients are referred with (normotensive) hypokalaemic alkalosis for investigation of a suspected SLT. We carried out a prospective, observational cohort study of patients referred to the UCL tubular clinic (the ‘London cohort’) between 2012 and 2017. The study was approved under the ethical approval ‘Identification of genes involved in renal, electrolyte and urinary tract disorders’ (REC reference 05/Q0508/6).

Patients enrolled were persistently hypokalaemic, requiring potassium supplementation. Patients were excluded if they were hypertensive, had Chronic Kidney disease (CKD stage 4/5), incomplete laboratory/clinical data or other causes for their hypokalaemia (e.g. familial hypokalaemic paralysis).

All London cohort patients were genotyped using the Multiplicom TUBMASTR panel by multiplex PCR and next generation sequencing, for a panel of genes associated with inherited tubular disease, including *SLC12A3, CLCNKB, HNF1B, SLC12A1, KCNJ1, BSND*^12^.

Contemporaneous data including demographics, blood and urine biochemistry, were collected from the electronic patient record. This included data for age, gender, blood pressure, serum and urinary biochemistry. From this the urinary fractional excretion (FE) of sodium, chloride, magnesium was calculated. The London cohort comprised 66 genotyped patients, with 31 datapoints per patient. Of these 39 had genetically confirmed SLT.

This data was analysed and descriptive statistics (including t-tests, Mann-Whitney tests and receiver operator characteristic curves) were generated using Graphpad PRISM and the open access statistics package R (https://www.r-project.org/), in order to identify variables that would be able to correctly classify patients as WT or SLT.

As similar studies have been published in the last 5 years, we decided to incorporate these data. A dataset from Taipei, Taiwan, published in 2017 (the ‘Taipei cohort’)^13^, comprised 87 patients, with 27 datapoints per patient. There were 43 genetically confirmed SLT patients in this cohort, who were selected in a very similar fashion to our own. A large Japanese cohort has also recently been published^14, 15^. This cohort (the ‘Kobe cohort’) was 266 patients strong. Each patient had 20 data points and there were 209 genotyped SLT patients.

As descriptive statistics had been published on these datasets already, we took the view that duplicating this would be redundant. We did, however, harmonise the datasets so that all they all had the same 18 datapoints and reanalysed the combined dataset using the same methods as the London cohort alone.

Additionally, we decided to employ an approach using machine learning (ML) algorithms to see whether we could correctly classify SLT patients on the basis of known clinical and biochemical datapoints in the whole dataset.

The London and Taipei cohorts were harmonised (to the common 18 datapoints present in all cohorts) and combined. This combined dataset was used as a ‘training dataset’ to train a number of ML algorithms.

Then, classification algorithms were applied to all 18 datapoints in the training dataset to classify the patients as WT or SLT. All of the algorithms were then rerun against the Kobe cohort which comprised the ‘testing dataset’.

The machine learning analyses were performed using the open access WEKA (Waikato Environment for Knowledge Analysis, version 3.8.3, University of Waikato, New Zealand) platform.

## Results

### London Cohort

The London cohort comprised 66 patients, ranging in age from 17 to 85 (mean age 41) years. (see Table 1). There were 38 female and 29 male patients.

**Table 1:** Bivariate analysis of the London cohort, Wildtype (WT) vs. Salt-losing tubulopathy (SLT) patients. Values displayed are median with interquartile range. Test shows which statistical test was used to determine significance, Mann Whitney (MW) for non-normally distributed data or the student’s T test (T-test) for normally distributed data.

|  | WT (n=27) | SLT (n=39) | Test | P-value | Diff in means (95% CI) |
| --- | --- | --- | --- | --- | --- |
| Age (years) | 42 (29-51) | 39 (28-45) | T-test | 0.05087 | 7.68 (-15.4-0.30) |
| Serum sodium (mmol/L) | 140 (139-141) | 141 (138-142) | T-test | 0.748 | 0.276 (-1.43-1.99) |
| Serum potassium (mmol/L) | 3.7 (3.4-4.1) | 3.2 (2.8-3.7) | T-test | 0.00479* | 0.49 (-0.817 - -0.153) |
| Serum urea (mmol/L) | 6.082 (3.725-7) | 6.113 (3.9-7.225) | T-test | 0.319 | 0.91 (-0.90-2.73) |
| Serum creatinine | 95.35 (72.25-110) | 76.47 (55.75-85.5) | MW | 0.6455 |  |
| Serum urate (mmol/L) | 0.3110 (0.2380-0.3900) | 0.271 (0.230-0.370) | MW | 0.4965 |  |
| Serum bicarbonate (mmol/L) | 28.85 (25-32) | 29.77 (27-31.75) | T-test | 0.41 | 1.10 (-1.62-3.85) |
| Serum chloride (mmol/L) | 97.9 (93.5-103) | 94.7 (92.5-98) | T-test | 0.20 | 2.17 (-5.43-1.18) |
| Urine calcium (mmol/L) | 1.800 (0.925-3.600) | 1.100 (0.50-2.50) | MW | 0.1468 |  |
| Serum corrected calcium (mmol/L) | 2.392 (2.305-2.48) | 2.406 (2.312-2.5) | T-test | 0.29 | 0.038 (-0.034-0.11) |
| Serum phosphate (mmol/L) | 1.168 (1.03-1.31) | 1.083 (1.002-1.188) | T-test | 0.24 | 0.67 (-0.18-0.046) |
| Serum magnesium (mmol/L) | 0.751 (0.638-0.853) | 0.662 (0.563-0.74) | T-test | 0.45 | 0.033 (-0.12-0.055) |
| Renin (nmol/L/hr) | 5.07 (1.76-5.26) | 11.71 (2.79-18.21) | MW | 0.00318* |  |
| Serum aldosterone (pmol/L) | 827 (242-1117) | 870 (210-930) | MW | 0.1868 |  |
| Urine sodium (mmol/L) | 68.1 (26.5-111) | 73.8 (36.8-91.3) | MW | 0.7263 |  |
| Urine potassium (mmol/L) | 74.72 (25-107.75) | 64.29 (45-78.75) | MW | 0.1416 |  |
| Urine creatinine (μmol/L) | 8380 (4305-11985) | 6000 (4045-7685) | MW | 0.00032* |  |
| Urine urea (mmol/L) | 198 (124-300) | 134 (83-175) | MW | 0.1971 |  |
| Urine chloride (mmol/L) | 46 (31.5-100.5) | 64.5 (41.5-108.25) | MW | 0.3576 |  |
| Urine magnesium (mmol/L) | 3.175 (1.175-4.525) | 2.095 (1.25-2.475) | MW | 0.02202* |  |
| Systolic blood pressure (mmHg) | 125 (113-134) | 116.9 (108.5-120) | MW | 0.4065 |  |
| Diastolic blood pressure (mmHg) | 78 (73-83) | 72.58 (70.5-74) | T-test | 0.26 | 5.42 (-15.68-4.79) |
| Weight (kg) | 78.0 (58.3-83) | 66.34 (56.7-79.4) | T-test | 0.78 | 2.73 (-22.4-16.9) |
| Fractional excretion of chloride (%) | 2.193 (1.145-3.283) | 2.353 (1.538-2.933) | T-test | 0.00069* | 0.73 (0.33-1.13) |
| Fractional excretion of sodium (%) | 0.587 (0.140-0.824) | 0.712 (0.387-1.00) | T-test | 0.0035* | 0.31 (0.11-0.52) |
| Fractional excretion of magnesium (%) | 4.92 (1.93-7.06) | 4.95 (2.29-9.12) | MW | 0.1645 |  |

Direct comparison between the SLT and WT groups shows significant differences for a number of variables. SLT patients tended to have lower serum potassium, magnesium, and higher serum renin concentrations. SLT patients had significantly greater polyuria, as evidenced by a lower urinary creatinine concentration. They had a higher fractional excretion of chloride and fractional excretion of sodium (see Table 1).

Receiver Operator Characteristic (ROC) curves were generated for the same data. The serum potassium was a fair discriminator of SLT vs WT (AUC 0.7 ± 0.07, p=0.007). The plasma renin concentration was a good discriminator (AUC 0.8 ± 0.07, p=0.003), as was the urinary creatinine concentration (AUC 0.8 ± 0.06, p<0.0001) see Figure 1.

**Figure 1:**
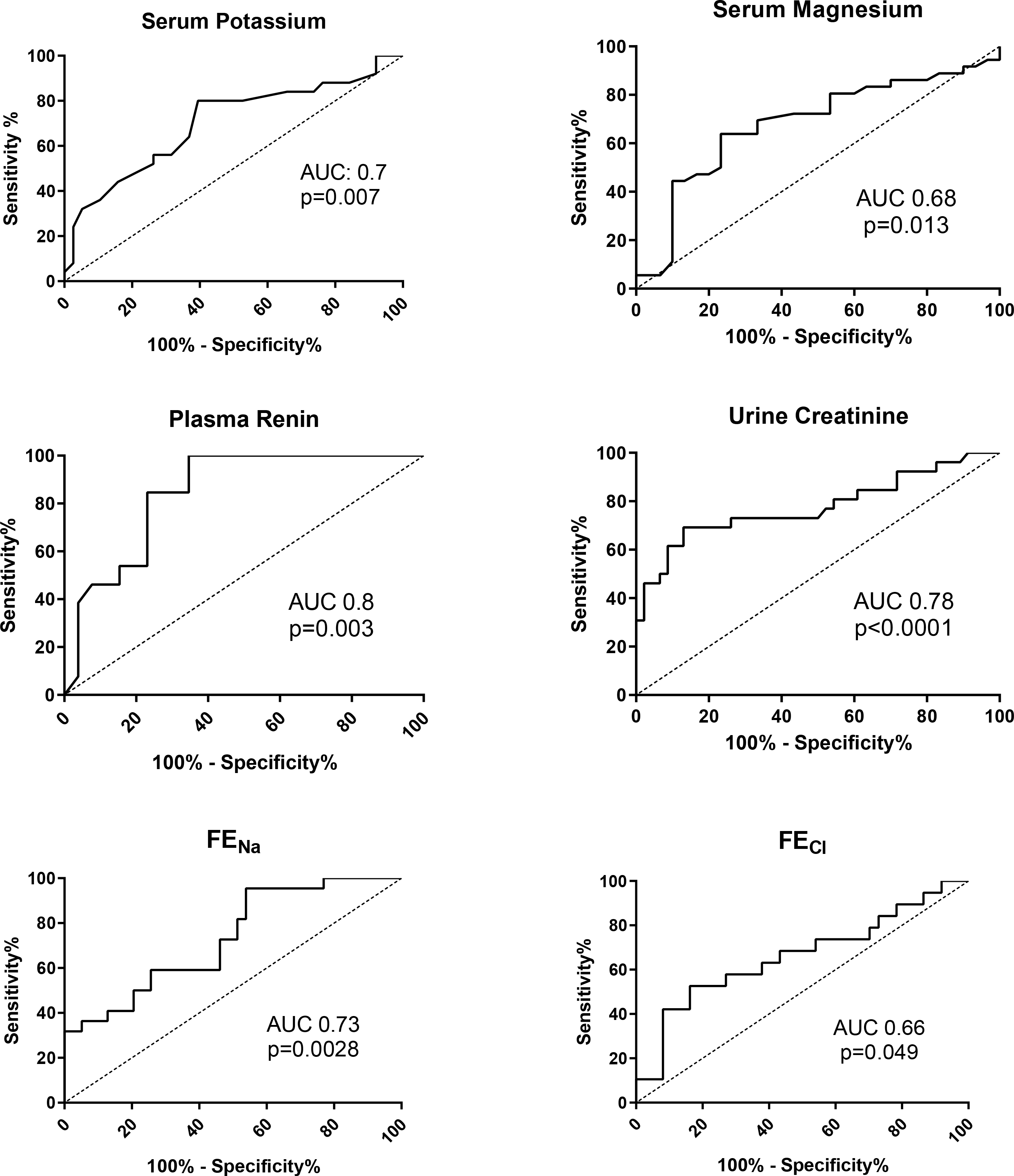
Receiver operator characteristic curves of significantly different biochemical values between SLT and WT in the London cohort. Area under the curve (AUC) and statistical significance (p value) indicated for each curve.

### Combined cohort

The combined data set (London, Taipei and Kobe cohorts) was 419 patients, of which 291 had genotyped SLT (see Figure 2). The median age was 26 (IQR 11-40) years. There were 239 female and 180 male patients. There were significant differences between the SLT and WT groups. SLT patients presented at a younger age. The serum potassium and aldosterone concentrations were lower in the SLT group. The urinary fractional excretion of sodium, chloride and magnesium were all significantly higher (all p<0.0001) in the SLT than the WT group (see Table 2).

**Figure 2:**
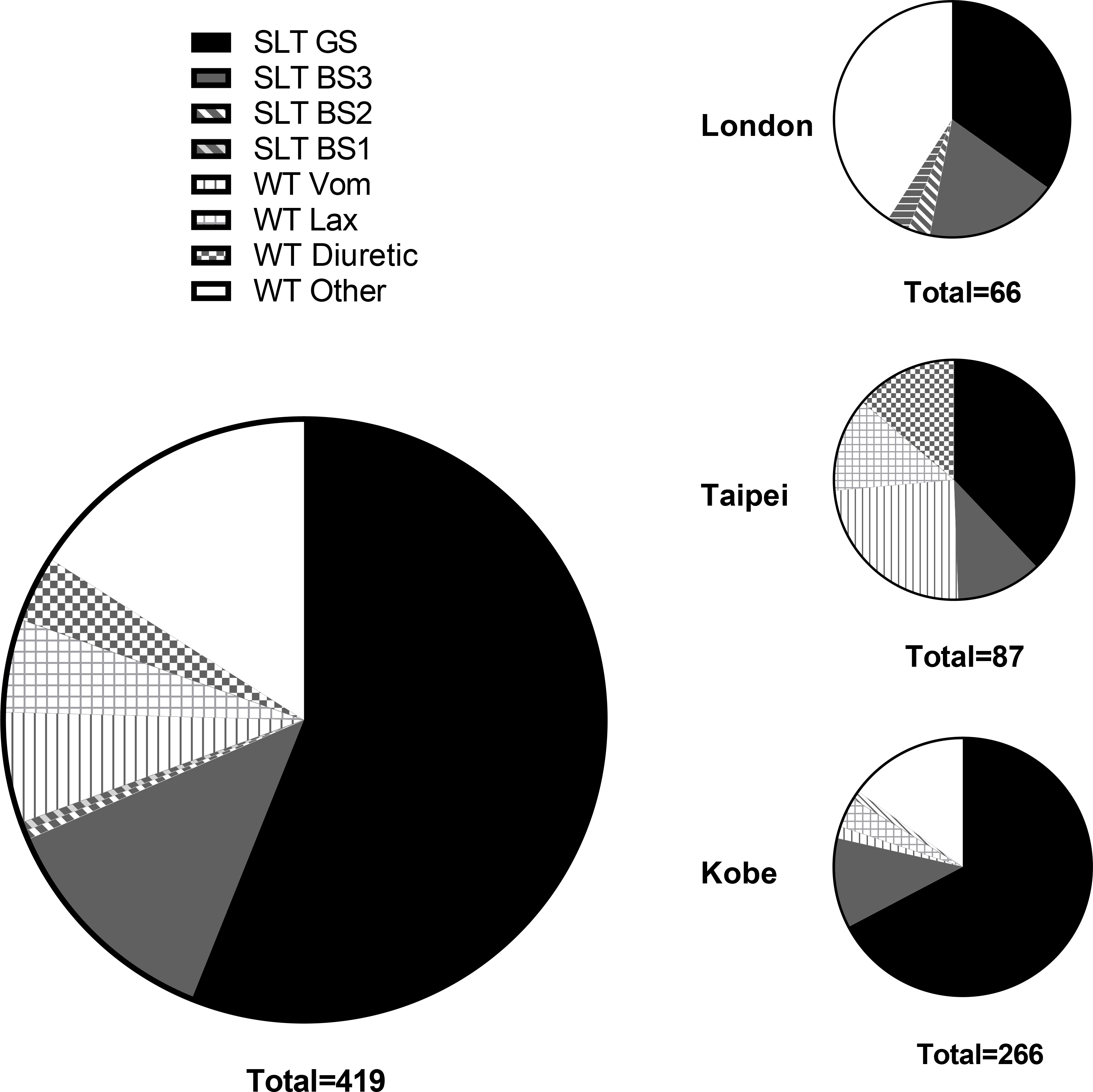
Proportion and subtype of SLT and WT patients in the London, Taipei, and Kobe cohorts (small figures) and the combined cohort (large figure).

**Table 2:**
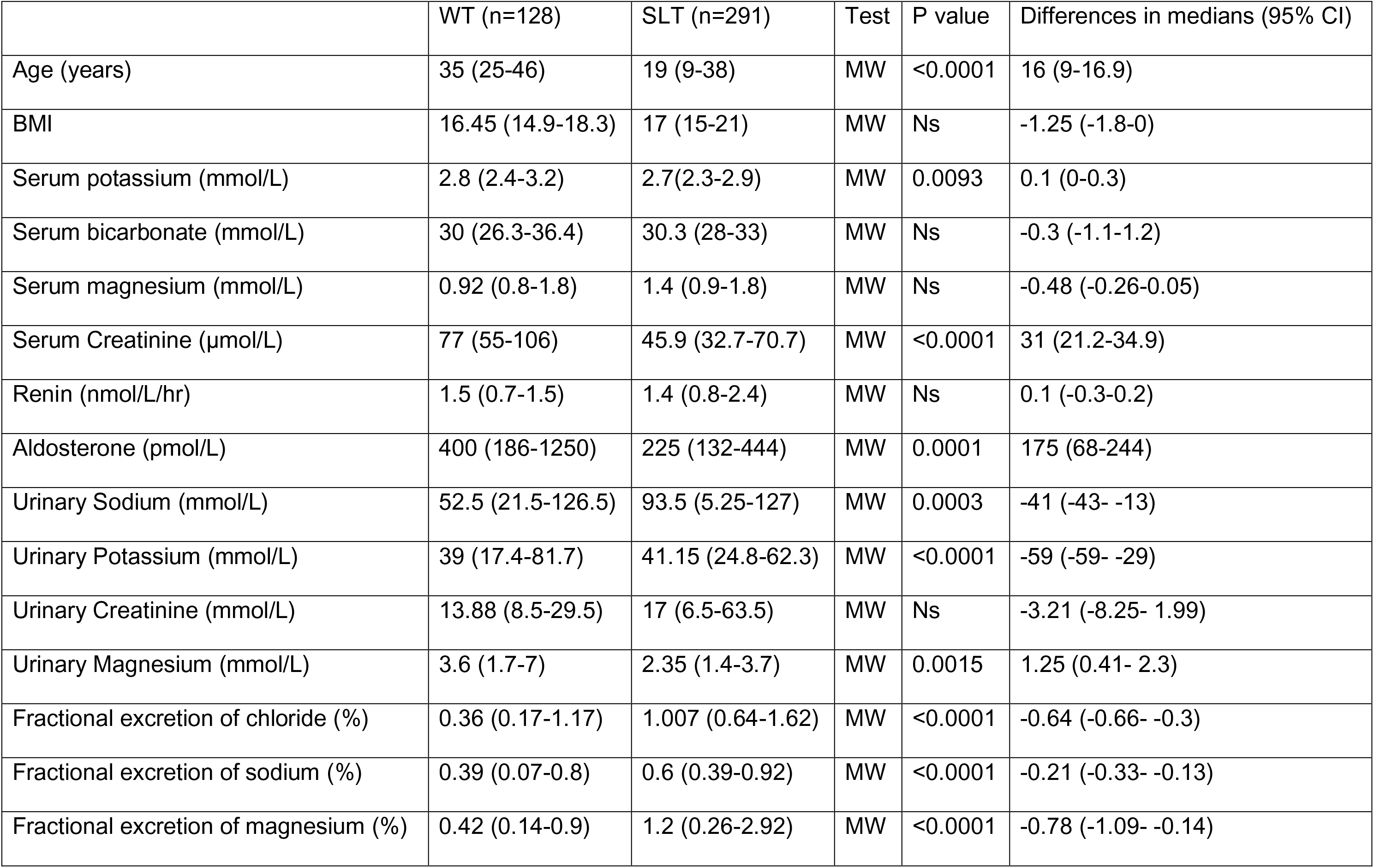
Bivariate analysis of the combined cohort, Wildtype (WT) vs. Salt-losing tubulopathy (SLT) patients Values displayed are median with interquartile range. Test shows which statistical test was used to determine significance, Mann Whitney (MW) for non-normally distributed data or the student’s T test (T-test) for normally distributed data.

**Table 3:** ML classification algorithm performance in differentiating Salt-losing tubulopathy (SLT) vs Wildtype (WT) patients, training dataset was the combined London and Taipei cohorts shaded rows), testing dataset was the Kobe cohort (clear rows). Measures of accuracy include the F-measure, Receiver Operator Characteristic area under the curve (ROC area) and Precision Recall Curve area under the curve (PRC area).

Generation of ROC curves showed that the urinary fractional excretion of chloride was a fair discriminator of SLT vs WT (AUC 0.69 ± 0.04, p<0.0001) and the fractional excretion of magnesium was a good discriminator (AUC 0.89 ± 0.03, p<0.0001). See Figure 3.

**Figure 3:**
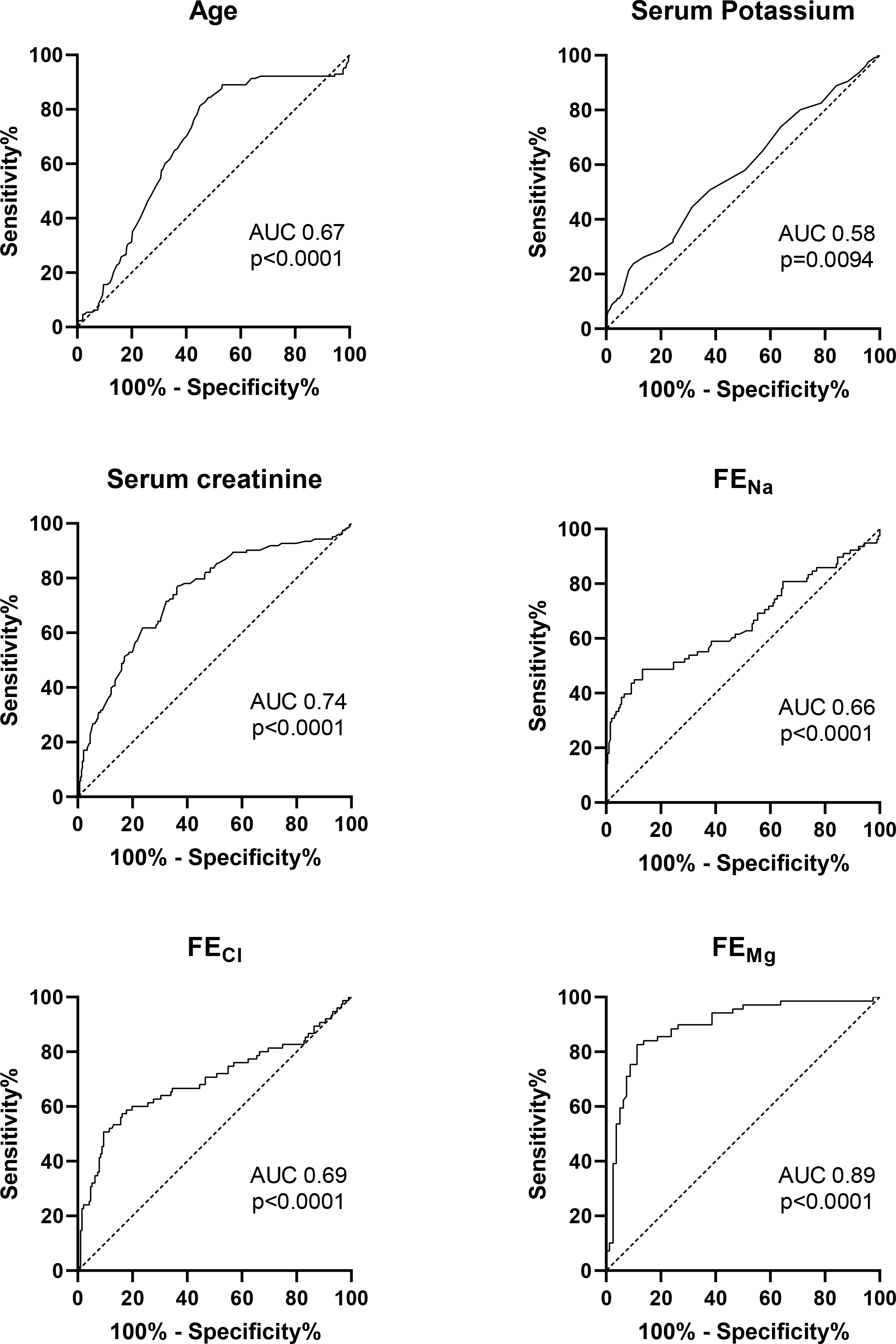
Receiver operator characteristic curves of significantly different biochemical values between SLT and WT in the combined cohort. Area under the curve (AUC) and statistical significance (p value) indicated for each curve.

### Machine Learning

Classification algorithms were run against the training set (London and Taipei cohorts), using 10-fold cross validation. These were then run against the testing dataset (Kobe cohort).

Intriguingly, the two algorithms with the highest accuracy in the testing set used simple rules to predict WT or SLT. These were a decision table and a ‘decision stump’ (an abbreviated decision tree with only one branch) algorithm. The better performing decision table (predictive accuracy in the training set 77% and 75% in the testing set) used a combination of the serum bicarbonate and the FE_Na_. The decision stump (predictive accuracy 66% and 61%) simply discriminated based on the FE_Cl_. These attributes were all predicted in the attribute selection step. These simple models are detailed in Figure 4.

**Figure 4:**
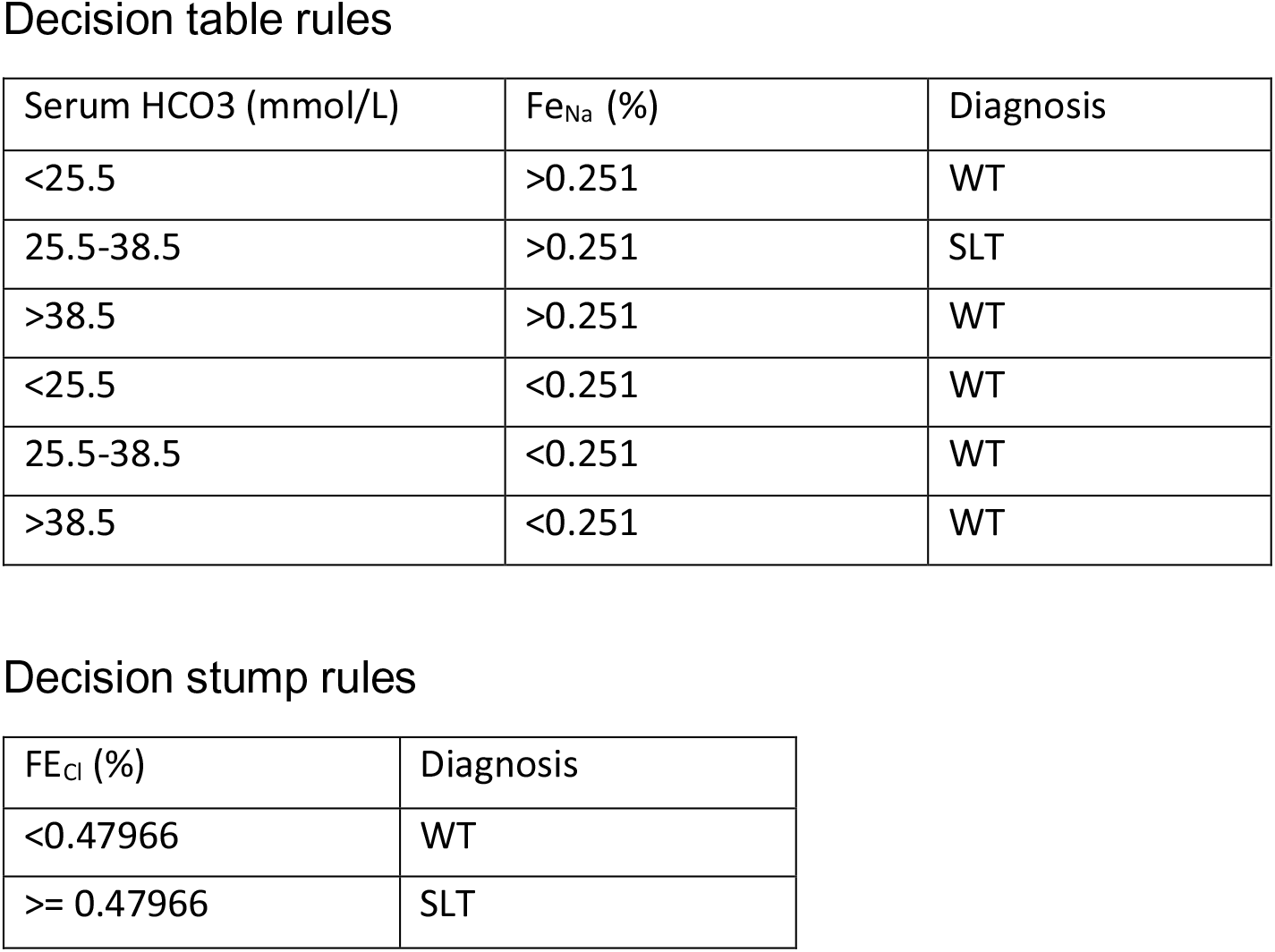
Simple rules used by the two most accurate classification algorithms to classify patients to a diagnosis of SLT or WT in the testing set. A) Rules for the Decision Table algorithm (predictive accuracy 75%) B) Rule for the Decision Stump algorithm (predictive accuracy 61%).

We compared these two algorithms to the single biochemical variables identified by conventional statistical methods as the two best discriminators in the London and combined cohorts. We did this by running each of these variables (FE_Mg_, renin, serum creatinine, urine creatinine) as a decision stump algorithm trained on the London and Taipei cohorts and tested on the Kobe cohort. The ML algorithms were superior discriminators than the variables identified by standard statistical methods in the London or combined cohorts (see Table 4), despite an apparently superior ROC AUC value for FE_Mg_ in the combined cohort.

**Table 4:** Comparison of the two best performing algorithms identified by ML (shaded columns) compared to the two best single biochemical variables identified by standard statistics in the London and combined cohorts (clear columns). Measures of accuracy include the F-measure, Receiver Operator Characteristic area under the curve (ROC area) and Precision Recall Curve area under the curve (PRC area).

|  | Decision Table<br>(FE<sub>Na</sub>/HCO<sub>3</sub>) | Decision Stump<br>(FE<sub>Cl</sub>) | Decision Stump<br>(FE<sub>Mg</sub>) | Decision Stump<br>(Serum Creatinine) | Decision Stump<br>(Renin) | Decision Stump<br>(Urinary Creatinine) |
| --- | --- | --- | --- | --- | --- | --- |
| Training set |  |  |  |  |  |  |
| Predictive accuracy | 77% | 66% | 64% | 73% | 58% | 63% |
| F-measure | 0.77 | 0.65 | 0.6 | 0.75 | 0.57 | 0.6 |
| ROC AUC | 0.79 | 0.69 | 0.58 | 0.7 | 0.52 | 0.6 |
| PRC AUC | 0.74 | 0.67 | 0.58 | 0.75 | 0.53 | 0.62 |
| Testing set |  |  |  |  |  |  |
| Predictive accuracy | 75% | 61% | 47% | 52% | 31% | 53% |
| F-measure | 0.73 | 0.62 | 0.52 | 0.49 | 0.2 | 0.57 |
| ROC AUC | 0.55 | 0.54 | 0.37 | 0.51 | 0.53 | 0.48 |
| PRC AUC | 0.7 | 0.7 | 0.64 | 0.51 | 0.67 | 0.66 |
| Rules |  |  |  |  |  |  |
| Predict WT | See Figure 4 | <0.48% | <0.52% | >54.5 | <=10.4 | >16.06 |
| Predict SLT |  | >=0.48% | >=0.52% | <=54.5 | >10.4 | <=16.06 |

As a secondary analysis we combined all of the cohorts into one harmonised dataset and retrained algorithms using 10-fold cross validation, using either the best performing 8 datapoints or all 18 (see table 5) This resulted in higher predictive accuracy, the best performing algorithm being a Random Forest (predictive accuracy 81.9%, ROC AUC 0.89).

**Table 5:** Secondary analysis of classification algorithm performance in differentiating Salt-losing tubulopathy (SLT) vs Wildtype (WT) patients using 10-fold cross validation on the entire combined cohort, using either all 18 attributes or the 8 most significant attributes selected in the attribute selection step. Measures of accuracy include the F-measure, Receiver Operator Characteristic area under the curve (ROC area) and Precision Recall Curve area under the curve (PRC area).

### Subgroup analyses

WT patients who had been assigned a diagnosis of vomiting or diuretic or laxative abuse (only the Taipei and Kobe cohorts) were compared. The most significant differences between the three groups were the urinary potassium, FE_Na_ and FE_Cl_ which are shown in Figure 5.

**Figure 5:**
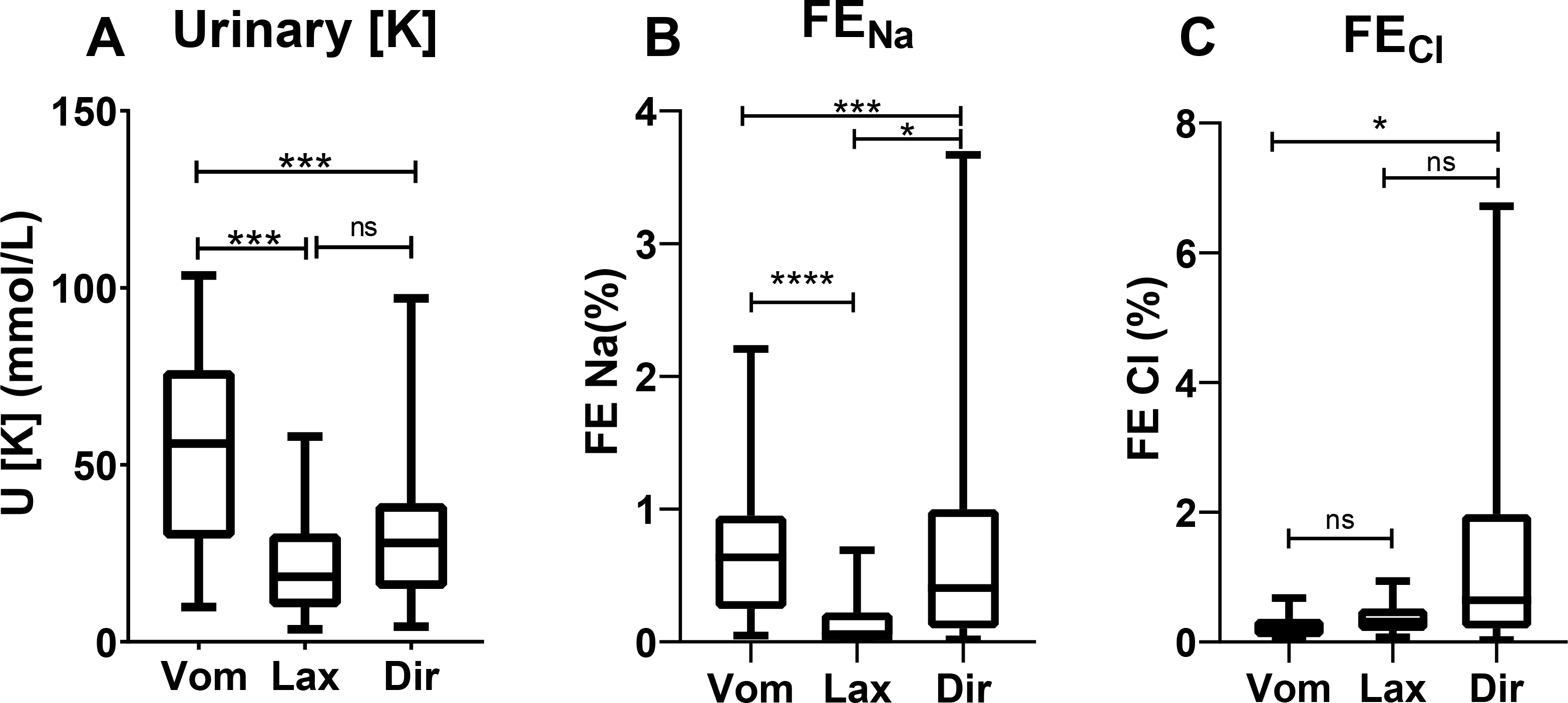
Significant biochemical differences between WT purging subgroups; vomitters (Vom), laxative abusers (Lax) and diuretic abusers (Dir). Significance is determined by the Mann Whitney test (between two groups) or the Krukshal-Wallis test (between all groups); ns=non-significant, *=p<0.05, ***=p<0.001, ****=p<0.0001

In the SLT patients, we also compared the GS patients to those with BS type 3. The most significant differences were in the serum bicarbonate, renin activity, aldosterone, urine creatinine, FE_Cl_ and FE_Mg_. These are shown in Figure 6.

**Figure 6:**
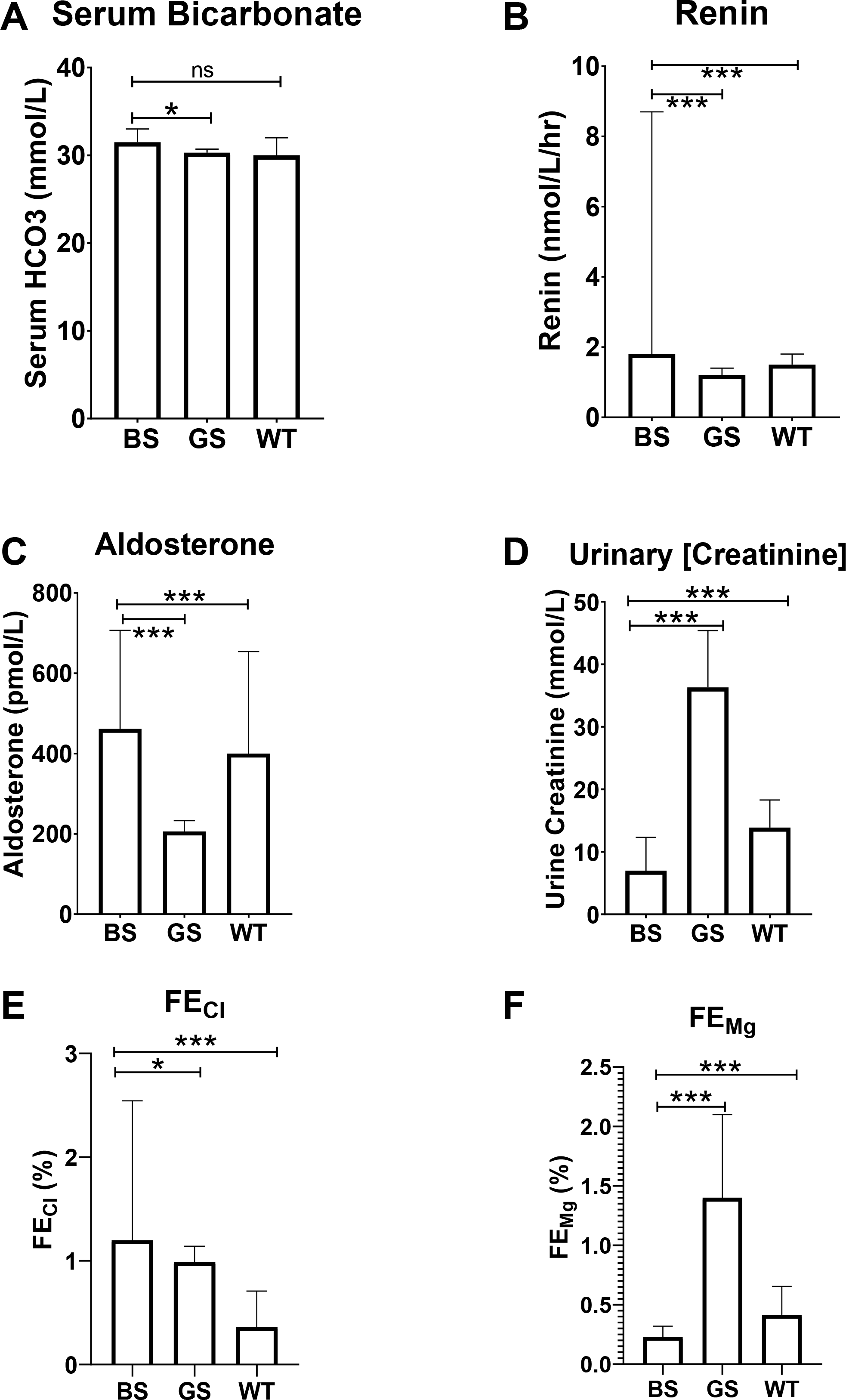
Significant biochemical differences between Type 3 Bartter syndrome (BS), Gitelman syndrome (GS) and wildtype (WT) patients in the combined cohort. Data expressed as medians with 95% confidence interval. Significance is determined by the Mann Whitney test (between two groups) or the Krukshal-Wallis test (between all groups); ns=non-significant, *=p<0.05, ***=p<0.001, ****=p<0.0001

### Genetic analysis

The genotyping results for SLT patients from the combined cohort are presented in table 6. There were 291 SLT patients, of which pathological variants were present in *SLC12A3* (235), *CLCNKB* (52), *SLC12A1* (2) and *KCNJ1* (2).

**Table 6:** Genotyping data from all SLT patients. The affected gene and mutation are detailed in columns 2-4. If a second mutation is present (i.e. a compound heterozygote), this is detailed in columns 5-6.

| Cohort | Gene | Mutation 1 | Mutation 1 (AA change) | Mutation 2 | Mutation 2 (AA change) |
| --- | --- | --- | --- | --- | --- |
| London | SLC12A3 | c.2891G>A | p.Asp311Glyfs*22 | c.2221G>A | p.Gly741Arg;p.Arg964Gln |
| London | SLC12A3 | c.1928C>T | p.Pro643Leu | Deletion exon 14 |  |
| London | SLC12A3 | c.1664C>T | p.Ser555Leu | c.2882+1G>T | p.(?) |
| London | SLC12A3 | c.1664C>T | p.Ser555Leu | c.2883+1G>T | p.(?) |
| London | SLC12A3 | c.427A>G | p.Met143Val | c.2221G>A | Gly741Arg |
| London | SLC12A3 | c.1028T>A | p.Met343Lys | c.1028T>A | p.Met343Lys |
| London | SLC12A3 | c.1000C>T | p.Arg334Trp | c.2221G>A | p.Gly741Arg |
| London | SLC12A3 | Deletion exons 1-7 |  | c.1825+1del | p.Glu609Argfs*2 |
| London | SLC12A3 | c.2186G>C | p.Gly729Ala | c.2883+1G>T | p.(?) |
| London | SLC12A3 | c.1939del | p.Val647CysfsTer25 | c.283+1G>T | p.(?) |
| London | SLC12A3 | c.1964G>A | p.Arg655His | c.2221G>A | p.Gly741Arg |
| London | SLC12A3 | c.910A>C | p.Thr304Pro | c.2883+1G>T | del exon 24 |
| London | SLC12A3 | c.111T>A | p.Tyr37Ter |  |  |
| London | SLC12A3 | c.1261T>C | p.Cys421Arg | c.1930del | p.Gln644SerfsTer |
| London | SLC12A3 | c.2221G>8 | Gly741Arg | c.2965G>A | Gly989Arg |
| London | SLC12A3 | c.237_238dup | p.Arg80ProfsTer35 | c.237_238dup | Frameshift starting Arg80 with new STOP codon 34 positions downstream |
| London | SLC12A3 | c.363G>C | p.Glu121Asp |  |  |
| London | SLC12A3 | c.1925G>A | p.Arg642His | c.2965G>A | Gly989Arg |
| London | SLC12A3 | c.2368+1del | p.(?) | SLC12A3_Ex6del |  |
| London | SLC12A3 | c.2576T>C | Leu859Pro | c.2965G>A | Gly989Arg |
| London | SLC12A3 | c.2221G>A | Gly741Arg | c.2581C>T* | Arg861Cys |
| London | SLC12A3 | c.961C>T | p.Arg321Trp | c.2883+1G>T | p.(?) |
| London | SLC12A1 | c.1215G>A | p.Glu405Glu possible splicing effect | c.1215G>A | p.Glu405Glu possible splicing effect |
| London | SLC12A1 | c.1316G>A | p.Arg439Gln | c.1316G>A | p.Arg439Gln |
| London | CLCNKB | Duplication Ex.4_8 |  |  |  |
| London | CLCNKB | c.1897del | p.Leu633Ter | c.1897del | p.Leu633Ter |
| London | CLCNKB | Whole gene deletion |  | Whole gene deletion |  |
| London | CLCNKB | Whole gene deletion |  | Whole gene deletion |  |
| London | CLCNKB | c.887G>A | p.Gly296Asp | c.1929+1G>A | p.(?) |
| London | CLCNKB | c.887G>A | p.Gly296Asp | c.1929+1G>A | p.(?) |
| London | CLCNKB | Whole gene deletion |  | Whole gene deletion |  |
| London | CLCNKB | c.610G>A | p.Ala204Thr | c.610G>A | p.Ala204Thr |
| London | CLCNKB | Whole gene deletion |  | Whole gene deletion |  |
| London | CLCNKB | c.1897delC | p.Leu633Ter |  |  |
| London | CLCNKB | c.1987A>T | p.Arg663Ter | c.1987A>T | p.Arg663Ter |
| London | CLCNKB | NC_000001.10:g.16383308_16383569del |  | g.16383308_16383569del |  |
| London | KCNJ1 | c.89G>A | p.Cys30Tyr |  |  |
| London | KCNJ1 | c.89G>A | p.Cys30Tyr | c.89G>A | p.Cys30Tyr |
| Kobe | CLCNKB | Ex1-2 deletion |  | c.1830G>A | p.Trp610* |
| Kobe | CLCNKB | Ex1-2 deletion |  | c.1830G>A | p.Trp610* |
| Kobe | CLCNKB | c.1830G>A | p.Trp610* |  |  |
| Kobe | CLCNKB | c.1830G>A | p.Trp610* |  |  |
| Kobe | CLCNKB | c.1830G>A | p.Trp610* |  |  |
| Kobe | CLCNKB | Ex1-2 deletion |  | c.1830G>A | p.Trp610* |
| Kobe | CLCNKB | Ex1-2 deletion |  | c.1830G>A | p.Trp610* |
| Kobe | CLCNKB | c.1830G>A | p.Trp610* |  |  |
| Kobe | CLCNKB | c.1334_1335delCT | p.Ser445Phefs*5 | c.1830G>A | p.Trp610* |
| Kobe | CLCNKB | c.1172G>A | p.Trp391* |  |  |
| Kobe | CLCNKB | Ex1-19 deletion |  | c.1830G>A | p.Trp610* |
| Kobe | CLCNKB | c.1172G>A | p.Trp391* |  |  |
| Kobe | CLCNKB | c.1830G>A | p.Trp610* |  |  |
| Kobe | CLCNKB | c.1830G>A | p.Trp610* |  |  |
| Kobe | CLCNKB | Ex1-19 deletion |  |  |  |
| Kobe | CLCNKB | c.1830G>A | p.Trp610* |  |  |
| Kobe | CLCNKB | c.1297+1G>A | IVS13+1G>A | c.1830G>A | p.Trp610* |
| Kobe | CLCNKB | c.100+2T>C | IVS2+2T>C | c.1830G>A | p.Trp610* |
| Kobe | CLCNKB | c.226C>T | p.Arg76* | c.1830G>A | p.Trp610* |
| Kobe | CLCNKB | c.1830G>A | p.Trp610* |  |  |
| Kobe | CLCNKB | c.1033G>A | p.Gly345Ser | c.1172G>A | p.Trp391* |
| Kobe | CLCNKB | c.1830G>A | p.Trp610* |  |  |
| Kobe | CLCNKB | c.910C>T | p.Arg304* | c.1830G>A | p.Trp610* |
| Kobe | CLCNKB | c.655G>T | p.Gly219Cys | c.1845+1G>A | IVS17ds+1 G>A |
| Kobe | CLCNKB | Ex15-19 duplication |  | c.1830G>A | p.Trp610* |
| Kobe | CLCNKB | c.1468G>A | p.Glu490Lys | Ex1-19 deletion |  |
| Kobe | CLCNKB | c.1830G>A | p.Trp610* |  |  |
| Kobe | CLCNKB | c.1309G>A | p.Gly437Arg | c.1830G>A | p.Trp610* |
| Kobe | CLCNKB | c.1309G>A | p.Gly437Arg | c.1830G>A | p.Trp610* |
| Kobe | CLCNKB | c.992_993insTCTGC | p.Thr332Leufs*19 | c.1172G>A | p.Trp391* |
| Kobe | SLC12A3 | c.1924C>T | p.Arg642Cys | c.2573T>A | p.Leu858His |
| Kobe | SLC12A3 | c.1195C>T | p.Arg399Cys | c.2573T>A | p.Leu858His |
| Kobe | SLC12A3 | c.1257_1259dup | p.Ala420dup | c.1868T>C | p.Leu623Pro |
| Kobe | SLC12A3 | c.2029G>A | p.Val677Met | c.2573T>A | p.Leu858His |
| Kobe | SLC12A3 | c.788_805dup18bp | p.Thr269delinsAsnTrpArgGlyLeuGlyPro | c.1196_1202dupGTGATGC | p.Ser402* |
| Kobe | SLC12A3 | c.1195C>T | p.Arg399Cys | c.1670-1G>T | IVS13 as G-T -1 |
| Kobe | SLC12A3 | c.788_805dup18bp | p.Thr269delinsAsnTrpArgGlyLeuGlyPro | c.3052C>T | p.Arg1018* |
| Kobe | SLC12A3 | c.1868T>C | p.Leu623Pro | c.2503C>T | p.Gln835* |
| Kobe | SLC12A3 | c.179C>T | p.Thr60Met | c.2029G>A | p.Val677Met |
| Kobe | SLC12A3 | c.139delC | p.His47Thrfs*67 | c.1924C>T | p.Arg642Cys |
| Kobe | SLC12A3 | c.2221G>A | p.Gly741Arg | c.2573T>A | p.Leu858His |
| Kobe | SLC12A3 | c.2573T>A | p.Leu858His |  |  |
| Kobe | SLC12A3 | c.1195C>T | p.Arg399Cys | c.1924C>T | p.Arg642Cys |
| Kobe | SLC12A3 | c.1195C>T | p.Arg399Cys | c.1924C>T | p.Arg642Cys |
| Kobe | SLC12A3 | c.2891G>A | p.Arg964Gln |  |  |
| Kobe | SLC12A3 | c.179C>T | p.Thr60Met |  |  |
| Kobe | SLC12A3 | c.788_805dup18bp | p.Thr269delinsAsnTrpArgGlyLeuGlyPro |  |  |
| Kobe | SLC12A3 | c.788_805dup18bp | p.Thr269delinsAsnTrpArgGlyLeuGlyPro | c.863T>C | p.Leu288Pro |
| Kobe | SLC12A3 | c.539C>A | p.Thr180Lys | c.1732G>A | p.Val578Met |
| Kobe | SLC12A3 | c.2573T>A | p.Leu858His | c.2891G>A | p.Arg964Gln |
| Kobe | SLC12A3 | c.1706C>T | p.Ala569Val | c.2573T>A | p.Leu858His |
| Kobe | SLC12A3 | c.139delC | p.His47Thrfs*67 | c.2573T>A | p.Leu858His |
| Kobe | SLC12A3 | c.2573T>A | p.Leu858His |  |  |
| Kobe | SLC12A3 | c.539C>A | p.Thr180Lys | c.1868T>C | p.Leu623Pro |
| Kobe | SLC12A3 | c.2573T>A | p.Leu858His | c.2747+2T>A | IVS23+2T>A |
| Kobe | SLC12A3 | c.2573T>A | p.Leu858His | c.2747+2T>A | IVS23+2T>A |
| Kobe | SLC12A3 | c.1262G>T | p.Cys421Phe |  |  |
| Kobe | SLC12A3 | c.539C>A | p.Thr180Lys |  |  |
| Kobe | SLC12A3 | c.539C>A | p.Thr180Lys |  |  |
| Kobe | SLC12A3 | c.817dupG | p.Ala273Glyfs*38 | c.1670-191C>T | IVS13 as C-T -191 |
| Kobe | SLC12A3 | c.1868T>C | p.Leu623Pro | c.2548+253C>T | IVS21 ds C-T +253 |
| Kobe | SLC12A3 | c.179C>T | p.Thr60Met | c.2573T>A | p.Leu858His |
| Kobe | SLC12A3 | c.1045C>T | p.Pro349Ser | c.1706C>T | p.Ala569Val |
| Kobe | SLC12A3 | c.539C>A | p.Thr180Lys | c.3053G>A | p.Arg1018Q |
| Kobe | SLC12A3 | c.960C>G | p.Tyr320* | c.2573T>A | p.Leu858His |
| Kobe | SLC12A3 | c.1100C>T | p.Pro367Leu | c.2573T>A | p.Leu858His |
| Kobe | SLC12A3 | c.1732G>A | p.Val578Met | c.2573T>A | p.Leu858His |
| Kobe | SLC12A3 | c.1844C>T | p.Ser615Leu | c.2537_2538delTT | p.Phe846* |
| Kobe | SLC12A3 | c.2573T>A | p.Leu858His | c.3052C>T | p.Arg1018* |
| Kobe | SLC12A3 | c.2573T>A | p.Leu858His | c.2877_2878delAG | p.Arg959Serfs*11 |
| Kobe | SLC12A3 | c.488C>T | p.Thr163Met | c.1939delG | p.Val647Cysfs*25 |
| Kobe | SLC12A3 | c.1195C>T | p.Arg399Cys | c.2891G>A | p.Arg964Gln |
| Kobe | SLC12A3 | c.539C>A | p.Thr180Lys | c.1195C>T | p.Arg399Cys |
| Kobe | SLC12A3 | c.179C>T | p.Thr60Met | c.1670-1G> T | IVS13 as G-T -1 |
| Kobe | SLC12A3 | c.539C>A | p.Thr180Lys | c.805_806ins18 | p.Thr269delinsAsnTrpArgGlyLeuGlyPro |
| Kobe | SLC12A3 | c.539C>A | p.Thr180Lys | c.1924C>T | p.Arg642Cys |
| Kobe | SLC12A3 | c.539C>A | p.Thr180Lys | c.2573T>A | p.Leu858His |
| Kobe | SLC12A3 | c.1924C>T | p.Arg642Cys | c.2029G>A | p.Val677Met |
| Kobe | SLC12A3 | c.2573T>A | p.Leu858His | c.2927C>T | p.Ser976Phe |
| Kobe | SLC12A3 | c.179C>T | p.Thr60Met | c.1670-1G> T | IVS13 as G-T -1 |
| Kobe | SLC12A3 | c.488C>T | p.Thr163Met | c.2573T>A | p.Leu858His |
| Kobe | SLC12A3 | c.179C>T | p.Thr60Met | c.3052C>T | p.Arg1018* |
| Kobe | SLC12A3 | c.1924C>T | p.Arg642Cys | c.2573T>A | p.Leu858His |
| Kobe | SLC12A3 | c.1706C>T | p.Ala569Val | c.2927C>T | p.Ser976Phe |
| Kobe | SLC12A3 | c.539C>A | p.Thr180Lys |  |  |
| Kobe | SLC12A3 | c.1924C>T | p.Arg642Cys | c.2573T>A | p.Leu858His |
| Kobe | SLC12A3 | c.2573T>A | p.Leu858His |  |  |
| Kobe | SLC12A3 | c.1868T>C | p.Leu623Pro | c.1963C>T | p.Arg655Cys |
| Kobe | SLC12A3 | c.506-1G>A | IVS3 as G-A -1 | c.2927C>T | p.Ser976Phe |
| Kobe | SLC12A3 | c.539C>A | p.Thr180Lys | c.1924C>T | p.Arg642Cys |
| Kobe | SLC12A3 | c.1216A>C | p.Asn406His | c.2927C>T | p.Ser976Phe |
| Kobe | SLC12A3 | c.817dupG | p.Ala273Glyfs*38 | c.1924C>T | p.Arg642Cys |
| Kobe | SLC12A3 | c.1868T>C | p.Leu623Pro | c.1930delC | p.Gln644Serfs*28 |
| Kobe | SLC12A3 | c.1868T>C | p.Leu623Pro | c.1930delC | p.Gln644Serfs*28 |
| Kobe | SLC12A3 | c.488C>T | p.Thr163Met | c.1963C>T | p.Arg655Cys |
| Kobe | SLC12A3 | c.3052C>T | p.Arg1018* |  |  |
| Kobe | SLC12A3 | c.505+5g>c | IVS3 ds G-C +5 | c.1868T>C | p.Leu623Pro |
| Kobe | SLC12A3 | c.178A>G | p.Thr60Ala | c.2573T>A | p.Leu858His |
| Kobe | SLC12A3 | c.178A>G | p.Thr60Ala | c.2573T>A | p.Leu858His |
| Kobe | SLC12A3 | c.664_666delATT | p.Ile222del | c.2573T>A | p.Leu858His |
| Kobe | SLC12A3 | c.1924C>T | p.Arg642Cys | c.2573T>A | p.Leu858His |
| Kobe | SLC12A3 | c.2537_2538delTT | p.Phe846* | c.2573T>A | p.Leu858His |
| Kobe | SLC12A3 | c.2537_2538delTT | p.Phe846* | c.2573T>A | p.Leu858His |
| Kobe | SLC12A3 | c.1868T>C | p.Leu623Pro |  |  |
| Kobe | SLC12A3 | c.539C>A | p.Thr180Lys | c.2573T>A | p.Leu858His |
| Kobe | SLC12A3 | c.1924C>T | p.Arg642Cys | c.1963C>T | p.Arg655Cys |
| Kobe | SLC12A3 | c.1924C>T | p.Arg642Cys | c.1963C>T | p.Arg655Cys |
| Kobe | SLC12A3 | c.1924C>T | p.Arg642Cys | c.2573T>A | p.Leu858His |
| Kobe | SLC12A3 | c.1930delC | p.Gln644Serfs*28 | c.2573T>A | p.Leu858His |
| Kobe | SLC12A3 | c.539C>A | p.Thr180Lys |  |  |
| Kobe | SLC12A3 | c.1924C>T | p.Arg642Cys | c.1930delC | p.Gln644Serfs*28 |
| Kobe | SLC12A3 | c.539C>A | p.Thr180Lys | c.1049C>T | p.Ser350Leu |
| Kobe | SLC12A3 | c.1963C>T | p.Arg655Cys | c.2927C>T | p.Ser976Phe |
| Kobe | SLC12A3 | c.1664C>T | p.Ser555Leu |  |  |
| Kobe | SLC12A3 | c.539C>A | p.Thr180Lys | c.668delT | p.Phe223Serfs*79 |
| Kobe | SLC12A3 | c.1670-1G>T | IVS13G>T | c.2573T>A | p.Leu858His |
| Kobe | SLC12A3 | c.1844C>T | p.Ser615Leu |  |  |
| Kobe | SLC12A3 | c.179C>T | p.Thr60Met | c.2927C>T | p.Ser976Phe |
| Kobe | SLC12A3 | c.788_805dup18bp | p.Thr269delinsAsnTrpArgGlyLeuGlyPro | c.3052C>T | p.Arg1018* |
| Kobe | SLC12A3 | c.788_805dup18bp | p.Thr269delinsAsnTrpArgGlyLeuGlyPro | c.3052C>T | p.Arg1018* |
| Kobe | SLC12A3 | c.817dupG | p.Ala273Glyfs*38 | c.3052C>T | p.Arg1018* |
| Kobe | SLC12A3 | c.2573T>A | p.Leu858His | c.3052C>T | p.Arg1018* |
| Kobe | SLC12A3 | c.2573T>A | p.Leu858His | c.3053G>A | p.R1018Q |
| Kobe | SLC12A3 | c.664_666delATT* | p.Ile222del | c.2573T>A | p.Leu858His |
| Kobe | SLC12A3 | c.1732G>A | p.Val578Met | c.2573T>A | p.Leu858His |
| Kobe | SLC12A3 | c.1A>T | p.Met1Leu | c.1868T>C p.L623P | p.Leu623Pro |
| Kobe | SLC12A3 | c.2573T>A | p.Leu858His | c.3052C>T | p.R1018* |
| Kobe | SLC12A3 | c.1924C>T | p.Arg642Cys | c.1930delC | p.Gln644Serfs*28 |
| Kobe | SLC12A3 | c.1278C>A* | p.Asn426Lys | c.1930delC | p.Gln644Serfs*28 |
| Kobe | SLC12A3 | c.488C>T | p.Thr163Met | c.2573T>A | p.Leu858His |
| Kobe | SLC12A3 | c.1844C>T | p.Ser615Leu | c.1930delC | p.Gln644Serfs*28 |
| Kobe | SLC12A3 | c.788_805dup18bp | p.Thr269delinsAsnTrpArgGlyLeuGlyPro | c.1868T>C | p.Leu623Pro |
| Kobe | SLC12A3 | c.2891G>A | p.Arg964Gln |  |  |
| Kobe | SLC12A3 | c.2573T>A | p.Leu858His |  |  |
| Kobe | SLC12A3 | c.2573T>A | p.Leu858His |  |  |
| Kobe | SLC12A3 | c.2573T>A | p.Leu858His | c.2891G>A | p.Arg964Gln |
| Kobe | SLC12A3 | c.539C>A | p.Thr180Lys | c.2573T>A | p.Leu858His |
| Kobe | SLC12A3 | c.1289G>A | p.Cys430Tyr | c.2573T>A | p.Leu858His |
| Kobe | SLC12A3 | c.788_805dup18bp | p.Thr269delinsAsnTrpArgGlyLeuGlyPro |  |  |
| Kobe | SLC12A3 | c.788_805dup18bp | p.Thr269delinsAsnTrpArgGlyLeuGlyPro |  |  |
| Kobe | SLC12A3 | c.788_805dup18bp | p.Thr269delinsAsnTrpArgGlyLeuGlyPro |  |  |
| Kobe | SLC12A3 | c.247C>T | p.Arg83Trp | c.2573T>A | p.Leu858His |
| Kobe | SLC12A3 | c.2537_2538delTT | p.Phe846* | c.2573T>A | p.Leu858His |
| Kobe | SLC12A3 | c.2029G>A | p.Val677Met | c.2573T>A | p.Leu858His |
| Kobe | SLC12A3 | c.2573T>A | p.Leu858His | c.3052C>T | p.R1018* |
| Kobe | SLC12A3 | c.2573T>A | p.Leu858His | c.2686C>T* | p.Arg896* |
| Kobe | SLC12A3 | c.1924C>T | p.Arg642Cys | c.2573T>A | p.Leu858His |
| Kobe | SLC12A3 | c.3052C>T | p.Arg1018* |  |  |
| Kobe | SLC12A3 | c.626G>C* | p.Arg209Pro | c.2029G>A | p.Val677Met |
| Kobe | SLC12A3 | c.1925G>A | p.Arg642His | c.3052C>T | p.Arg1018* |
| Kobe | SLC12A3 | c.2573T>A | p.Leu858His | c.2891G>A | p.Arg964Gln |
| Kobe | SLC12A3 | c.788_805dup18bp | p.Thr269delinsAsnTrpArgGlyLeuGlyPro |  |  |
| Kobe | SLC12A3 | c.539C>A | p.Thr180Lys | c.2573T>A | p.Leu858His |
| Kobe | SLC12A3 | c.1732G>A | p.Val578Met | c.2573T>A | p.Leu858His |
| Kobe | SLC12A3 | c.1271G>A* | p.Gly424Asp |  |  |
| Kobe | SLC12A3 | c.704C>T* | p.Thr235Met | c.1709C>T | p.Ala570Val |
| Kobe | SLC12A3 | c1201_1210delCCTCTGGGG | p.Ala401_Gly403del | c.2891G>A | p.Arg964Gln |
| Kobe | SLC12A3 | c.911C>T | p.Thr304Met | c.2573T>A | p.Leu858His |
| Kobe | SLC12A3 | c.788_805dup18bp | p.Thr269delinsAsnTrpArgGlyLeuGlyPro | c.1289G>A | p.Cys430Tyr |
| Kobe | SLC12A3 | c.788_805dup18bp | p.Thr269delinsAsnTrpArgGlyLeuGlyPro | c.1289G>A | p.Cys430Tyr |
| Kobe | SLC12A3 | c.788_805dup18bp | p.Thr269delinsAsnTrpArgGlyLeuGlyPro | c.1826-1G>A | IVS14-1G>A |
| Kobe | SLC12A3 | c.2573T>A | p.Leu858His | c.3052C>T | p.Arg1018* |
| Kobe | SLC12A3 | c.1456G>A | p.Asp486Asn | c.1868T>C | p.Leu623Pro |
| Kobe | SLC12A3 | c.1924C>T | p.Arg642Cys | c.2686C>T | p.Arg896* |
| Kobe | SLC12A3 | c.1924C>T | p.Arg642Cys | c.2573T>A | p.Leu858His |
| Kobe | SLC12A3 | c.1A>T | p.Met1Leu | c.179C>T | p.Thr60Met |
| Kobe | SLC12A3 | c.2537_2538delTT | p.Phe846* | c.1923C>G | p.Tyr641* |
| Kobe | SLC12A3 | c.1077C>G | p.Asn359Lys | c.1709C>T | p.Ala570Val |
| Kobe | SLC12A3 | c.1868T>C | p.Leu623Pro | c.2927C>T | p.Ser976Phe |
| Kobe | SLC12A3 | c.2891G>A | p.Arg964Gln | ex9-18deletion |  |
| Kobe | SLC12A3 | c.2573T>A | p.Leu858His | ex7-8deletion |  |
| Kobe | SLC12A3 | c.1924C>T | p.Arg642Cys | c.3053G>A | p.Arg1018Q |
| Kobe | SLC12A3 | c.1930delC | p.Gln644Serfs*28 | c.2573T>A | p.Leu858His |
| Kobe | SLC12A3 | c.1930delC | p.Gln644Serfs*28 |  |  |
| Kobe | SLC12A3 | c.1963C>T | p.Arg655Cys | c.2573T>A | p.Leu858His |
| Kobe | SLC12A3 | c1924C>T | p.Arg642Cys | c.2573T>A | p.Leu858His |
| Kobe | SLC12A3 | c.539C>A | p.Thr180Lys | c.1698C>A | p.Asn566Lys |
| Kobe | SLC12A3 | c179C>T | p.Thr60Met | c.2099T>C | p.Leu700pro |
| Kobe | SLC12A3 | c.1924C>T | p.Arg642Cys | c.2573T>A | p.Leu858His |
| Kobe | SLC12A3 | c.238delC | p.(Arg80Glyfs*34) | c.2573T>A | p.Leu858His |
| Kobe | SLC12A3 | c.2573T>A | p.Leu858His | c.2891G>A | p.Arg964Gln |
| Kobe | SLC12A3 | c.539C>A | p.Thr180Lys | c.1868T>C | p.Leu623Pro |
| Kobe | SLC12A3 | c.1669+297T>G | P.(?) | c.2927C>T | p.Ser976Phe |
| Kobe | SLC12A3 | c.1897dupG | p.E633Gfs56 | c.1924C>T | p.Arg642Cys |
| Kobe | SLC12A3 | c.2191G>A | p.Gly731Arg |  |  |
| Kobe | SLC12A3 | c.1456G>A | p.Asp486Asn | c.2927C>T | p.Ser976Phe |
| Kobe | SLC12A3 | c.179C>T | p.Thr60met | c.2573T>A | p.Leu858His |
| Kobe | SLC12A3 | c.1100C>T | p.Pro367Leu | c.1924C>T | p.Arg642Cys |
| Kobe | SLC12A3 | c.2573T>A | p.Leu858His | c.2573T>A | p.Leu858His |
| Kobe | SLC12A3 | c.539C>A | p.Thr180Lys | c.2573T>A | p.Leu858His |
| Kobe | SLC12A3 | c.539C>A | p.Thr180Lys | c.1868T>C | p.Leu623Pro |
| Kobe | SLC12A3 | c.539C>A | p.Thr180Lys | c.2891G>A | p.Arg964Gln |
| Kobe | SLC12A3 | c.2573T>A | p.Leu858His | c.2573T>A | p.Leu858His |
| Kobe | SLC12A3 | c.2573T>A | p.Leu858His | c.3052C>T | p.Arg1018* |
| Kobe | SLC12A3 | c.1924C>T | p.Leu642Cys | c.2573T>A | p.Leu858His |
| Kobe | SLC12A3 | c.2573T>A | p.Leu858His | c.2573T>A | p.Leu858His |
| Kobe | SLC12A3 | c.1942C>T | p.Leu642Cys | c.2573T>A | p.Leu858His |
| Kobe | SLC12A3 | c.2927G>T | p.Ser976Phe | c.2927G>T | p.Ser976Phe |
| Kobe | SLC12A3 | c.2573T>A | p.Leu858His | c.2573T>A | p.Leu858His |
| Kobe | SLC12A3 | c.1868T>C | p.Leu623Pro | c.2927C>T | p.Ser976Phe |
| Kobe | SLC12A3 | c.1195C>T | Arg399Cys | c.2573T>A | p.Leu858His |
| Kobe | SLC12A3 | c.2573T>A | p.Leu858His | c.3053G>A | p.Arg1018Gln |
| Kobe | SLC12A3 | c.788_805dup18bp | p.Thr269delinsAsnTrpArgGlyLeuGlyPro | c.1132G>A | p.Ala378Thr |
| Kobe | SLC12A3 | c.1100C>T | p.Pro367Leu | c.1942C>T | p.Leu642Cys |
| Kobe | SLC12A3 | c.2573T>A | p.Leu858His |  |  |
| Kobe | SLC12A3 | c.1163C>A | p.Ala388Asp | c.2573T>A | p.Leu858His |
| Kobe | SLC12A3 | c.139delC | p.His47Thrfs*67 | c.2660+1G>A |  |
| Kobe | SLC12A3 | c.1163C>A | p.Ala388Asp | c.2573T>A | p.Leu858His |
| Kobe | SLC12A3 | c.539C>A | p.Thr180Lys | c.788_805dup18bp | p.Thr269delinsAsnTrpArgGlyLeuGlyPro |
| Taipei | SLC12A3 | c.1670-191C>T | 238bp-containing pseudo-exon | c.1670-191C>T | 238bp-containing pseudo-exon |
| Taipei | SLC12A3 | c.268C>T | p.His90Tyr | c.268C>T | p.His90Tyr |
| Taipei | SLC12A3 | c.37G>C/c.179C>T | p.Ala13Pro/p.Thr60Met | c.248G>A | p.Arg83Gln |
| Taipei | SLC12A3 | c.2129C>A | p.Ser710X | c.2875-2876delAG | p.Arg959fs |
| Taipei | SLC12A3 | c.248G>A | p.Arg83Gln | c.1670-191C>T | 238bp-containing pseudo-exon |
| Taipei | SLC12A3 | c.911C>T | p.Thr304Met | c.2875-2876delAG | p.Arg959fs |
| Taipei | SLC12A3 | c.1000C>T | p.Arg334Trp | c.1326C>G | p.Asn442Lys |
| Taipei | SLC12A3 | c.2875-2876delAG | p.Arg959fs |  |  |
| Taipei | SLC12A3 | c.179C>T | p.Thr60Met | c.179C>T | p.Thr60Met |
| Taipei | SLC12A3 | c.2875-2876delAG | p.Arg959fs | c.2875-2876delAG | p.Arg959fs |
| Taipei | SLC12A3 | c.2875-2876delAG | p.Arg959fs | c.2875-2876delAG | p.Arg959fs |
| Taipei | SLC12A3 | c.185A>G / c.248G>A | p.Asp62Gly/ p.Arg83Gln | c.1541C>T | p.Ala514Val |
| Taipei | SLC12A3 | c.179C>T | p.Thr60Met | c.2573T>A | p.Leu858His |
| Taipei | SLC12A3 | c.2542G>A | p.Asp848Asn |  |  |
| Taipei | SLC12A3 | c.2542G>A | p.Asp848Asn |  |  |
| Taipei | SLC12A3 | c.1946C>T | p.Thr649Met |  |  |
| Taipei | SLC12A3 | c.1946C>T | p.Thr649Met |  |  |
| Taipei | SLC12A3 | IVS7-1G>A (correct: c.577-1G>A)+c.965-976<br>GCGGACATTTTGT><br>ACCGAAAATTTT | p.(?); probably Exon7 and 8 skipping | c.1924C>T | p.Arg642Cys |
| Taipei | SLC12A3 | c.1670-191C>T | 238bp-containing pseudo-exon | Ex23 deletion |  |
| Taipei | SLC12A3 | c.488C>T /c.2612G>A | p.Thr163Met/p.Arg871His | c.2929C>A | p.Arg977X |
| Taipei | SLC12A3 | c.488C>T /c.2612G>A | p.Thr163Met/p.Arg871His | c.2929C>A | p.Arg977X |
| Taipei | SLC12A3 | c.1670-191C>T | 238bp-containing pseudo-exon | c.1670-191C>T | 238bp-containing pseudo-exon |
| Taipei | SLC12A3 | c.1670-191C>T | 238bp-containing pseudo-exon | c.1670-191C>T | 238bp-containing pseudo-exon |
| Taipei | SLC12A3 | c.2532G>A | p.Trp844X | c.2875-2876delAG | p.Arg959fs |
| Taipei | SLC12A3 | c.488C>T /c.2612G>A | p.Thr163Met/p.Arg871His | c.2129C>A | p.Ser710X |
| Taipei | SLC12A3 | c.488C>T /c.2612G>A | p.Thr163Met/p.Arg871His | c.2129C>A | p.Ser710X |
| Taipei | SLC12A3 | c.2129C>A | p.Ser710X | c.2548+253C>T | 90bp-containing pseudo-exon |
| Taipei | SLC12A3 | IVS7-1G>A+c.965-976<br>GCGGACATTTTGT><br>ACCGAAAATTTT | Exon7 and 8 skipping | c.2875-2876delAG | p.Arg959fs |
| Taipei | SLC12A3 | c.179C>T | p.Thr60Met | c.185A>G | p.Asp62Gly |
| Taipei | SLC12A3 | c.2129C>A | p.Ser710X |  |  |
| Taipei | SLC12A3 | c.2129C>A | p.Ser710X | c.2875-2876delAG | p.Arg959fs |
| Taipei | SLC12A3 | c.488C>T /c.2612G>A | p.Thr163Met/p.Arg871His | c.1278C>A | p.Asn426Lys |
| Taipei | SLC12A3 | c.488C>T /c.2612G>A | p.Thr163Met/p.Arg871His | c.2548+253C>T | 90bp-containing pseudo-exon |
| Taipei | SLC12A3 | c.1967C>T | p.P656L |  |  |
| Taipei | CLCNKb | c.752delG | p.L252Sfs | Ex1-19deletion |  |
| Taipei | CLCNKb | c.1004T>C | p.Lys335Pro | c.1409G>A | p.Gly470Glu |
| Taipei | SLC12A1 | c.825T>A/c.1493C>T | p.Tyr275X/p.Ala498Val | c.2996G>A | p.Arg999His |

## Discussion

Inherited salt-wasting disorders are uncommon and can pose a diagnostic and therapeutic challenge^16^. Presentation is typically with the (often incidental) finding of hypokalaemia, in the setting of a variable metabolic alkalosis without hypertension.

In adults, differential diagnoses are unlikely to include other known genetic diseases with the exception of *HNF1B* mutations, which in the early stages may phenocopy GS/BS3. Disorders such as congenital chloride diarrhoea and cystic fibrosis present in infancy and are not viable differential diagnoses in adults. The same does not hold true for novel genes causing SLT, or SLT-like syndromes.

Thus, the main differential diagnoses in adults comprise patients with eating disorders engaging in purging behaviours like vomiting, surreptitious diuretic or laxative use^8^. The eating disorders associated with purging are prevalent; anorexia nervosa has a lifetime prevalence of up to 2.2%^17^; bulimia has a lifetime prevalence of 1%^18^. The prevalence of purging behaviour within these eating disorders is as high as 86%^19^. This is a much higher prevalence than Gitelman syndrome, the most prevalent of the SLTs.

Biochemical clues to patients engaging in purging behaviour have been described. Prolonged vomiting will cause a metabolic alkalosis, which may be very severe^20, 21^; indeed it has been reported that a serum bicarbonate of >45 mmol/L is almost always due to a gastric cause^22^. Hypokalaemia is due to urinary wasting (from both hyperaldosteronism (from volume contraction) and bicarbonaturia (from acid loss); the latter may be more important as hypokalaemia may be ameliorated by proton pump inhibition^23^). Avid chloride retention typically results in undetectably low urinary chloride concentrations.

Chronic diarrhoea in laxative abusers causes an alkalosis in part from volume depletion and hyperaldosteronism, this is rarely severe; serum bicarbonate is rarely >34mmol/L^24^. Potassium losses are intestinal, and urinary potassium and chloride concentrations are low.

Surreptitious diuretic use will cause a hypokalaemic metabolic alkalosis. However, there will be a chloruresis while the drug is working, this will resolve when the drug effect ceases. This variability in the urinary chloride concentration may help to make the diagnosis^25^, which can be confirmed by detection of diuretic in the urine^26^.

However, differentiating individual purging behaviours may be difficult, or even futile. Purging patients are unlikely to admit these behaviours even on close questioning^27^. Furthermore, purging patients rarely employ only one purging behaviour; multiple purging behaviours are reported in as much as 52.5% of patients with eating disorders associated with purging^19^.

Therefore, we decided against trying to differentiate WT patients on the basis of their probable (or, occasionally proven) purging behaviour, and treated them as one homogenous group in the London cohort, and in the overall analysis.

The 2017 consensus report from KDIGO recommends that “genetic testing should be offered to all patients with a clinical suspicion of Gitelman Syndrome (minimal criteria)”^11^. However, access to nationally accredited methods to make a genetic diagnosis of Bartter or Gitelman syndrome are not universally available.

Moreover, genetic screening has some technical difficulties. Historically, apparent heterozygosity for SLT causing genes is common in patients presenting as SLT, as high as 15-20%^28, 29^ However, the apparent lack of pathogenic variants on the other allele may have been due to large genomic rearrangements that are missed by direct sequencing and may account for up to 15% of the genetic defects responsible for autosomal recessive diseases^30^.

The frequency for heterozygote carriage of pathogenic mutations in SLC12A3 is as high as 1-2.9%^31^. SLC12A3 heterozygosity is often reported in patients presenting with hypokalaemia, and as far as blood pressure goes they may have an intermediate phenotype between normal and Gitelman syndrome^32^. The significance of this observation is not known.

Furthermore, due to the distribution of CLCKb (in the TAL and the early DCT), there is considerable overlap in the phenotype of Type 3 Bartter and Gitelman syndrome, which may present identically^33, 34^. Seyberth proposed a different nomenclature to better reflect the correlation of the clinical features with the location in the nephron of the genetic lesion^6^. Phenotypic overlap also exists with *HNF1B* (which encodes the transcription factor HNF-1β); patients may present with typical electrolyte disturbances seen in GS, especially hypomagnesaemia and hypocalciuria^35^. While to date, most centres offering genetic testing for GS would routinely also sequence *CLCNKB*, *HNF1B* is not normally also sequenced, although it was as part of the TUBMASTR panel used for the London cohort.

Finally, and rather obviously, routine genetic screening will not pick up patients who have pathogenic variants in novel genes that cause a salt-losing nephropathy. In this sense, the KDIGO recommendation that genetic screening should be a mandatory investigation for these patients is germane, especially if it is in the context of a panel of genes that include other diseases that may phenocopy GS, such as *HNF1B*. It seems likely that whole exome or genome sequencing will replace single or multiple gene sequencing, which may make the detection of variants in putative novel genes possible.

In the London cohort, one of the best discriminators between SLT and WT patients is the plasma renin activity. RAAS activation is expected in SLT^36^, but would be expected in purging patients also; it may be that intermittent volume depletion is a lesser renin stimulus than constant renal salt loss.

A raised fractional excretion of chloride (FE_Cl_) has previously been reported in the SLT population^37^ and has been suggested as a part of clinical diagnostic criteria for Gitelman syndrome^11^. In our cohort we observed a statistically significant difference between the FE_Cl_ and FE_Na_ in both groups (each higher in SLT than WT). We also found that the urinary creatinine concentration was significantly lower in SLT; we interpreted this as indicating greater polyuria in SLT compared to WT.

We found a lower serum potassium level in SLT compared to WT patients at their first clinic visit (difference in means 0.49, 95% CI -0.817 to -0.153, p = 0.00479). This probably reflects the median difference between SLT patients with a constantly low serum potassium, compared to WT patients with a variable hypokalaemia from purging^38^.

In the combined cohort, age at presentation was significantly younger in SLT than WT; this is probably due to the young age of presentation in SLT, particularly in the Kobe cohort^14^. As in the London cohort, serum potassium was significantly lower in SLT, although this was a poor discriminator. The urinary fractional excretion of sodium, chloride and magnesium were all significantly increased in SLT, with the FE_Mg_ being the best discriminator.

However, the machine learning algorithms that performed best in both the training and testing datasets were a decision table that used the serum bicarbonate and FE_Na_ and a decision stump that only used the FE_Cl_. The decision table warrants some comment; it predicts SLT only when there is a relative natriuresis (FE_Na_>0.251%) and the serum bicarbonate is between 25.5 and 38.5 mmol/L; while those with the same sodium excretion and a more severe alkalosis (serum bicarbonate >38.5 mmol/L) were predicted as WT. Although very severe alkaloses have occasionally been reported in SLT, this in the more severe infantile forms of BS^39^; in adults, severe alkaloses is most commonly associated with persistent vomiting^20, 21^. The relative naturesis in the same WT patients is counterintuitive; one would expect a low FE_Na_ in a vomiting patient. Yet in our subgroup analysis, the WT patients deemed to be vomiting had a median FE_Na_ of 0.64% (IQR 0.25-0.95%). This is hard to explain unless there is more than one purging behaviour, e.g. diuretic abuse or using salt as an emetic.

The less accurate decision stump used only the FE_Cl_, in keeping with previous observations^8^, in fact a recent series of SLT diagnosed in childhood reported a median FE_Cl_ of 1.62%^40^. The combined cohort SLT patients had a median FE_Cl_ of 1.01% (IQR 0.64-1.67%) compared to WT who had a low median FE_Cl_ of 0.35% but a large IQR (0.17-1.17%), likely due to the contribution of diuretic-induced chloruresis in diuretic-abusing patients.

The predictive accuracy of these ML classification algorithms was better than the individual biochemical variables identified by standard statistical methods. Interestingly, it was also superior to that of expert clinicians; a recent report of the results of genotyping renal tubular disorders in adults in three European reference centres found a genetic confirmation rate of only 46% for Gitelman syndrome^12^. Lastly, we considered all patients with GS or type 3 BS to be a homogeneous group, ‘SLT’. Clinically, there can be significant overlap between the two groups, and GS was and (often still is) considered a mild adult form of BS. Indeed, Graziani stated that “the differential diagnosis between Gitelman syndrome and Bartter syndrome is more a semantic than a clinical issue”^36^.

## Conclusions

We present a new cohort of patients that have been investigated for SLT, all of whom have been comprehensively genotyped for genes known to cause or phenocopy SLT. We also present a combined dataset of this and two other, recently published cohorts in order to further interrogate the biochemical differences between SLT and WT patients. We present ML generated biochemical strategies to discriminate between SLT and WT, based on the largest combined cohort of genotyped SLT patients described, to our knowledge, in the literature. This data demonstrates the difficulties faced in trying to make a diagnosis based on biochemistry alone and underscores the necessity for comprehensive genetic diagnosis in this difficult set of patients.

### Author Contributions

EW and SBW conceived the study. DI and EA processed genetic data. CCS, HSL, CN and KN compiled data. DB, KS, BM and SBW analysed data. EW, DB, KS and SBW wrote the manuscript.

## Data Availability

All data produced in the present study are available upon reasonable request to the authors

## Supplementary Methods

ROC vs Predictive accuracy/Precision recall:

There is another factor to bear in mind when considering the different performances of the machine learning algorithms and the single variables selected by standard statistical methods. It is easy to see that the combined dataset that we have used is highly skewed toward patients with genetic SLT (see Figure 2), which comprise >50% of all of the cohorts in the dataset, and 67% of the combined dataset.

In such skewed datasets, a precision recall curve will often give a better idea of the performance of an algorithm than the ROC curve^41^. This may be a problem if the skewed nature of the dataset is unrepresentative of datasets that the algorithms will be used on. Obviously, this is not true in the general population and we would not argue that these algorithms are useful for population-based screening for SLTs. However, in hypokalaemic patients in whom a diagnosis of SLT is queried, the proportion of actual SLT is likely to be very high, and thus our datasets are likely representative of the relevant clinical scenarios.

